# Feasibility of Low-Cost Virtual Reality Motor Amplification for Stroke Rehabilitation

**DOI:** 10.1101/2024.08.23.24312233

**Authors:** Aidan Fisk, Summer Fox, Jenna Floyd, Christine Ausman, Daniel Blustein

## Abstract

**Purpose:** Virtual reality (VR) shows promise for supporting stroke rehabilitation and motor recovery. One design feature is motor amplification, a reinforcement-based approach with early evidence for reducing learned non-use among stroke survivors. However, its clinical use remains limited, with little understanding of its technical feasibility and user experience. To inform the ongoing development of VR rehabilitation systems, we assessed the technical feasibility and tolerability of automated, controller-free motor amplification in healthy young adults.

**Design:** Here we outline the initial development of the REVIVE system, a VR stroke rehabilitation system operating on the standalone Meta Quest line of Head Mounted Displays which are wireless, low cost, and require no external hardware. Hand tracking, voice recognition and an automated motor amplification algorithm enable accessible engagement for users. An animated coach in the virtual environment guides users through gamified exercises which simulate activities of daily living and functional movements. We tested the system with 60 healthy young adults, with a primary goal of validating the feasibility of the controller-free amplification feature on a low-cost headset.

**Findings:** Users reported minimal visually induced motion sickness even when experiencing the visuomotor perturbation generated by amplification, and more positive attitudes toward VR technology after the experience. Additionally, we provide a young healthy reference dataset for several REVIVE tasks to serve as a healthy baseline for future research with clinical populations.

**Value:** Our findings suggest that the addition of motor amplification to a consumer-grade system is technically feasible, has good user acceptance, and demonstrates short-term tolerability.

---

Worldwide, stroke ranks among the top causes of disability (IHME 2024b). Increasing survival rates over the past 30 years due to improved treatments have led to lower death rates but more years lived with disability (IHME 2024a), indicating a growing demand for post-stroke rehabilitation care. But patients typically receive less care than they need (Stockley *et al*., 2019).

Access to care is a barrier to maximal recovery, with barriers that include cost, distance to treatment centers, and limited transportation (Shanmugasegaram *et al*., 2013). These accessibility barriers are particularly acute in rural settings and in developing nations. In Canada, rural patients were less likely to receive stroke unit care (36% versus 51%), to see a neurologist (26% versus 65%), or to see an occupational therapist (52% versus 72%) compared to urban patients (Koifman *et al*., 2016). This mirrors treatment disparity globally with patients in Low- or Middle-Income Countries (LMICs) being less likely to receive standard care recommendations (Kayola *et al*., 2023; Owolabi *et al*., 2021).

Increasing access to intensive post-stroke rehabilitation therapies is an important goal that could have a substantial impact on patient outcomes worldwide. Telehealth and other technology-enabled solutions can help overcome these barriers to treatment and have been shown to be as effective and cost-efficient as in-person stroke care (Caughlin *et al*., 2020), enabling an increase in therapeutic dosing and access (“Canadian Stroke Best Practice Recommendations”, 2022). Public acceptance of telehealth solutions is high (“Canadian Stroke Best Practice Recommendations”, 2022), including among adults 65 years and older who make up about 75% of stroke cases (Yousufuddin and Young, 2019). Limitations persist as technology-enabled stroke rehabilitation options can place additional time and training burdens on clinicians (Cormican *et al*., 2023), be costly (Krakauer *et al*., 2021) and/or require multi-component hardware setups (Tsekleves *et al*., 2016; Uswatte *et al*., 2021). Addressing these limitations requires rehabilitation technologies to be designed around end-users in early design phases, emphasizing user-informed development to improve usability, adherence, and future clinical adoption (Broderick *et al*., 2023; Mitchell *et al*., 2023).

Efforts to use low-cost VR headsets, such as the Meta Quest 2, to deliver stroke rehabilitation have demonstrated beneficial impacts on stroke recovery (Amin *et al*., 2024; Herrera *et al*., 2023). Some evidence suggests that immersive VR confers greater benefit than conventional rehabilitation in measures of shoulder, wrist and hand function (Kiper *et al*., 2023), and gross motor function (Soleimani *et al*., 2024). Conventional rehabilitation, such as constraint-induced movement therapy (CIMT) targets learned non-use by restraining the unaffected limb and forcing repeated use of the paretic limb (Kwakkel *et al*., 2015; Teasell *et al*., 2020). While effective, CIMT’s reliance on physical restraint and high-intensity supervised practice limits its accessibility outside specialized clinical settings (Viana and Teasell, 2012). Whereas VR allows for dynamic adjustments to task parameters, such as shifting interactable objects closer to a user with a limited range of motion, these systems can adapt to the different abilities of users (Bressler *et al*., 2024; Herrera *et al*., 2024). These approaches typically rely on changing the environment, whereas an alternative approach, motor amplification, amplifies the user’s movements to reduce their motor deficit in the virtual world. Preliminary research has demonstrated that motor amplification can reduce paretic limb non-use in stroke patients by temporarily reducing post-stroke motor deficits, facilitating task completion and reducing frustration, thus leveraging the power of reinforcement learning (Ballester *et al*., 2015, 2016). Motor amplification confers advantages over environmental manipulation. First, studies have demonstrated that performing hand movements with augmented visual feedback in VR can enhance motor recovery and stimulate neural reorganization in stroke survivors (Adamovich *et al*., 2009; Bagce *et al*., 2012; Merians *et al*., 2009). Second, by modifying one’s movement as opposed to their environment, users are more likely to associate their successful reach completion with their own physical effort, which can lead to greater agency and body ownership compared to what results with environmental manipulations (Nataraj *et al*., 2020).

Attempts to implement amplification on low-cost headsets have been limited, with one requiring the need for controllers and manual setting of parameters (“WalkinVR”, n.d.), reducing its ease of use. While the benefits of amplification have been established in the lab using controller-based systems (Ballester *et al*., 2015; Wang *et al*., 2020), realizing the accessibility benefit of low-cost VR requires the dedicated validation of automated amplification on a controller-free, low-cost platform. Additionally, the user acceptance of amplification, which intentionally introduces a visuomotor perturbation, is less understood and requires validation.

The introduction of a visuomotor mismatch makes the assessment of visually induced motion sickness (VIMS) a critical component of this validation. Commonly explained by sensory conflict theory (LaViola, 2000; Oman, 1990), VIMS represents a mismatch between visual and vestibular or proprioceptive cues, just as amplification in VR introduces, and could be a barrier to adoption. Although low-cost VR rehabilitation systems have shown high levels of engagement and satisfaction (Mc Kittrick *et al*., 2023), further insight into the user experience with movement amplification is needed to determine the approach’s feasibility and usability. We therefore focused on measuring VIMS levels and user acceptance of the technology as primary outcomes of this validation.

To fill a need for more affordable and user-friendly stroke rehabilitation options, we developed the REVIVE virtual reality system (Fisk et al., 2025) (Fig. 1). The REVIVE system (<u>R</u>ehabilitation and <u>E</u>ngagement through <u>V</u>irtual <u>I</u>mmersi<u>v</u>e <u>E</u>nvironments) uses a head-mounted display and an animated virtual coach to guide patients through exercises, reducing the need for constant clinician presence. Avatar-guided and virtual-coaching approaches have been shown to improve exercise adherence and engagement in VR rehabilitation contexts (Han *et al*., 2026; Pourghasem *et al*., 2026). Features include gamified tasks for high-dose motor exercise, hand tracking to eliminate the need for handheld controllers, and wireless operation. The REVIVE software runs on a commercially available headset, the Meta Quest 2/3S/3, which costs about the same as four physiotherapy sessions, enabling widespread deployment.

**Figure 1.**
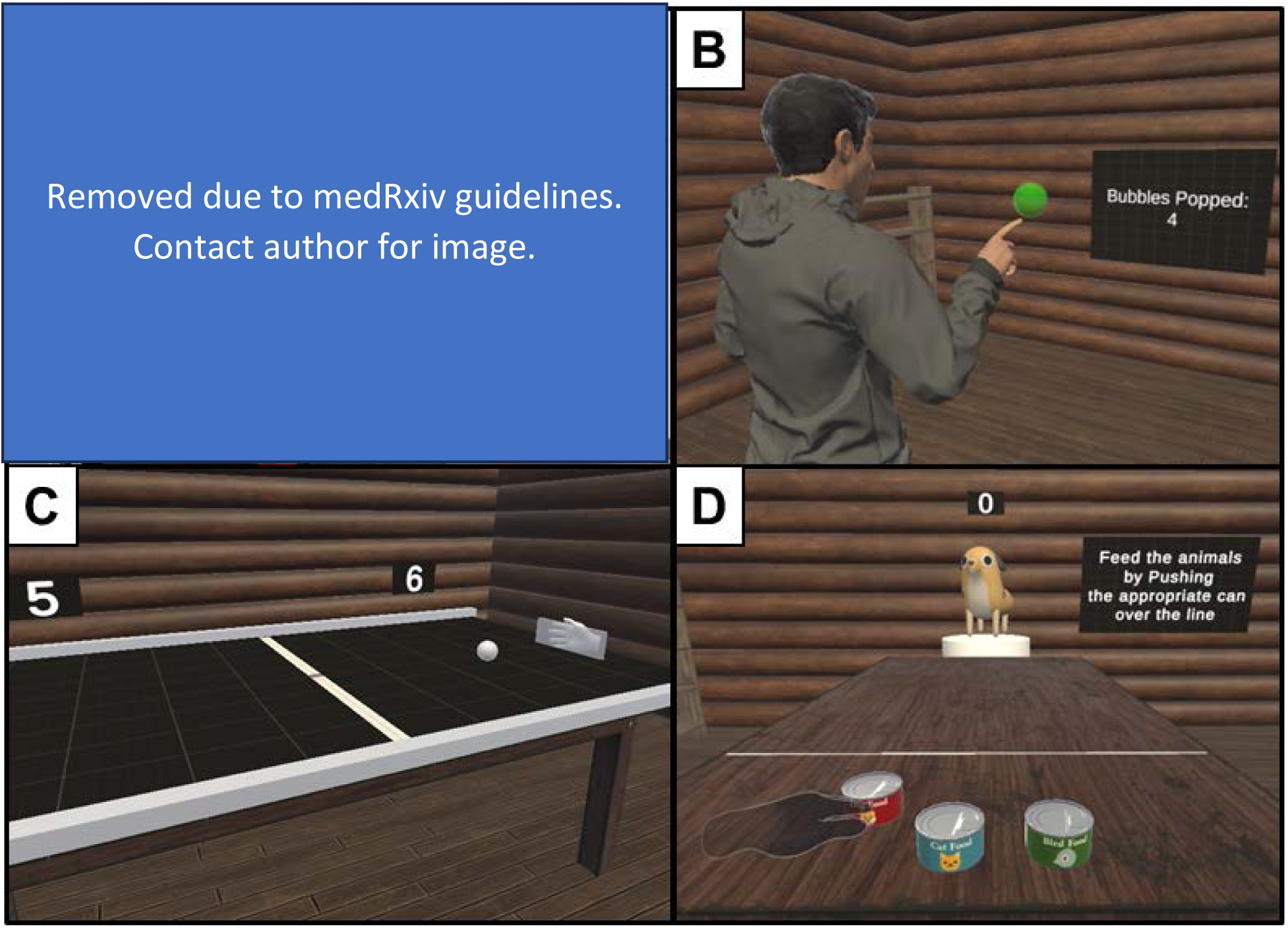
REVIVE system overview. A. User interacting with virtual environment through a Meta Quest 2 head mounted display. B. User view within the virtual environment during the Bubble Pop Game. C. User view during the Pong Game. D. User view during the Animal Feeding Task.

This paper presents the methodological development and initial proof-of-concept validation of the REVIVE system. We assessed the feasibility of automated, controller-free motor amplification in a sample of 60 young adults without disability and generated a young healthy reference dataset that can be used to make clinical comparisons in future studies. The primary focus of the feasibility testing was to determine participant receptiveness to the technology as measured by a VR attitudes survey before and after the simulated rehabilitation session. This emphasis on user attitudes and tolerability reflects a patient-centered approach to technology validation, in which acceptability is treated as a prerequisite to efficacy testing rather than a secondary outcome (Cornet *et al*., 2019; Perski and Short, 2021). Demonstrating the technology as accessible is critical to increasing its acceptance by patients, particularly in individuals who may have less experience with emerging technologies such as older adults (Ortiz-Fernandez *et al*., 2021). We also elaborate on the prevalence of VIMS (Fisk et al., 2025), which could be a barrier to the adoption of VR-based stroke rehabilitation. Our findings will inform the ongoing development of the REVIVE system and contribute to the ongoing discourse related to low-cost rehabilitation options for stroke, particularly in LMICs and rural areas with reduced access to care (Kayola *et al*., 2023; Owolabi *et al*., 2021).

## Methods

### REVIVE System

We created a software program featuring 12 gamified tasks, with three tested in this study. These tasks were designed to either simulate daily activities, like pouring water into a glass, or to act as immersive games where the hand acts as a controller in such classic games as Pong or Snake. Each task targeted one of three movement categories: 1) Gross reaching; 2) Reach and grasp; or 3) Precision grasping.

In this study, the software was run on the Meta Quest 2 head mounted display, an immersive virtual reality system that can operate wirelessly (the software also runs similarly on the more recently released Meta Quest 3S and 3). Onboard cameras track the hands of the user, allowing the user to interact with objects in the virtual space without the need for handheld controllers or external camera setups. Real-world physics are simulated in the virtual environment using the Unity Game Engine. Some object interactions behave differently from their real-world analogue, such as grasping an object, and these incongruencies are demonstrated and explained in an initial tutorial for first-time users. We did not run the interaction tutorial in this study since the tasks completed in this study involved only gross reaching movements.

#### Motor Amplification

At the beginning of each session, the user completes a guided calibration protocol which measures the range of motion (ROM) of each arm and hand. This protocol involved a bimanual reaching and gasping task in which participants were guided to reach towards a red box and grasp their hands. The hand-tracking system would record the participant’s ROM within a 3D space wherein the x-axis (left-right), y-axis (up-down), and z-axis (forward-backward) were all measured in meters. The bilateral difference in ROM was used to identify the ‘deficient’ limb that would be amplified in the virtual world to match the ROM of the ‘unaffected’ limb. There was no set minimum or maximum amplification as the amount of amplification was determined through the calibration in relation to the ‘unaffected’ limb, and amplification was applied in all directions (as opposed to dimension-specific). We applied a motor smoothing algorithm (Unity Technologies, 2026) to reduce the jitteriness induced by amplified hand tracking noise, and smoothing was proportional to the amount of amplification applied.

Hand-tracking errors would lead to a momentary disappearance of the virtual hand. If hand-tracking was not working at the start of the session, the system would be restarted. There were no participant reports of hand-tracking errors.

### Usability Study

Following the initial development of the REVIVE system (Fisk et al., 2025), we conducted early-stage proof-of-concept feasibility testing with healthy young adult participants. This study was approved by the Acadia University Research Ethics Board (REB File #22-07). Sixty students from Acadia University receiving course credit for their involvement were recruited for the study. As an initial proof-of-concept and feasibility study, this sample size (30 per group) was chosen to provide sufficient data to establish young healthy reference dataset and detect potential usability issues, consistent with similar early-stage technology assessments. Participants provided demographic data, including gender identity with option to self-identity, handedness, and age.

#### Study Protocol

This double-blind study used block randomization to assign participants to either the motor amplification group or the control group with no amplification. Participants donned a Meta Quest 2 VR headset connected to a desktop computer that ran the VR exercises. The experimenter monitored the virtual environment displayed to the participants and offered verbal guidance as necessary. The VR session lasted 10 to 15 minutes and included an initial calibration phase to assess participants’ range of motion. Surveys were administered to collect data on participants’ attitudes toward VR (pre- and post-session), prior technology use (pre-session), and VIMS (post-session).

#### VR Task Overview

Participants began with a calibration task to establish their range of motion, after which motor amplification was applied for the experimental group. They then performed three tasks: a Bubble Pop game, a Pong game, and an Animal Feeding task. The order of the last two tasks was counterbalanced, with half of the participants in each group completing the Pong game first and the other half beginning with the Animal Feeding task. These tasks were developed in consultation with clinicians working in the Acute Stroke Unit at Valley Regional Hospital (Kentville, NS, Canada), including an occupational therapist, a physiotherapist, a nurse practitioner, and a stroke unit coordinator (nurse working in an administrative role). Clinician involvement included initial discussions and iterative design with feedback sessions where clinicians tested the system and provided feedback. A description of each task is provided below:

##### Calibration Task

Participants reached for colored cubes positioned at different locations in their environment. The software recorded the range of motion for each hand and calculated the amplification factor for the experimental group based on the proportional difference between the two sides. This information was also used to set interaction boundaries for the subsequent tasks. Outcome measures included the volume of the ROM envelope and the amplification factor applied.

##### Bubble Pop Game

Participants popped virtual bubbles appearing in their field of view over a one-minute period. Hand positions were tracked at a frequency of 50 Hz to record movement trajectories. Outcome measures included the total number of bubbles popped, the overall distance traveled by the hands, and the 95th percentile hand velocity.

##### Pong Game

Participants controlled a virtual paddle by moving their hand forward or backward. The hand with the smaller range of motion was the controlled hand, with amplification applied in the experimental condition. A ball rolled across the table, and participants earned points by successfully deflecting the ball past the computer-controlled paddle. After a one-minute practice round, participants played a two-minute game. Outcome measures included the number of paddle-ball contacts, points scored, and opponent points scored.

##### Animal Feeding Task

Participants sat at a table in front of three cans of animal food arranged in a line from left to right. Each can was adorned with the image of an animal (dog, cat, bird) to indicate which animal’s food it contained. A virtual animal appeared at the far end of the table, and participants were required to push the matching can across a designated line away from the participant to “feed” the animal. The line’s distance was calibrated to the individual’s range of motion. Once the task was completed, the animal changed appearance and emitted an identifying sound (e.g. a bark for the dog), prompting the participant to push a new can across the line. Participants practiced for one minute before completing a two-minute session. The outcome measure was the number of correctly pushed cans.

#### Surveys

After providing written informed consent, participants completed surveys that collected demographic information and pre-intervention data on VR experience and attitudes. One question asked, “How often have you used virtual reality systems?”, with the options to select being *Very frequently*, *Frequently*, *Occasionally*, *Rarely*, *Never*. In the VR attitudes survey (Table I), participants responded to six statements using a Likert-scale [*Strongly Agree* (4), *Agree* (3), *Neutral* (2), *Disagree* (1), *Strongly Disagree* (0)]. The numerical equivalent scores reported for Statements 2 through 5 (Table I) were summed and their mean was used as an overall metric indicating attitudes towards VR technology. Scores for Statements 3 and 6 were reverse-coded.

**Table I.** Statements used to assess attitudes towards virtual reality with Likert-scale responses.

| Statement Number | Statement |
| --- | --- |
| 1 | I am very comfortable using technology (e.g., cellphones, computers, gaming systems). |
| 2 | I am very comfortable in virtual environments that are created using virtual reality systems. |
| 3 | I am concerned that I could damage the virtual reality system when I use it. |
| 4 | I think that virtual reality systems are easy to use. |
| 5 | I think that virtual reality could cause mental fatigue. |
| 6 | I am motivated to learn how to use virtual reality systems. |

Following the VR exercises, an abbreviated VIMS symptom assessment was administered, and the VR attitudes survey was readministered. The difference between the pre-survey and post-survey mean scores was used to indicate the overall change in VR attitude attributed to the VR experience. The VIMS symptom assessment consisted of 5 of 9 items from the Virtual Reality Sickness Questionnaire (VRSQ) (Kim *et al*., 2018): General Discomfort, Fatigue, Eyestrain, Difficulty Focusing, Headache. The five items were selected to minimize survey burden while capturing the most common and relevant symptoms of VIMS for a short VR session. We also sought to reduce redundancy across survey items and focus on aspects most likely to affect stationary VR users. The mean score across the five items was used to indicate VIMS immediately following the VR experience.

### Analysis Plan

Range of motion and motor performance scores on each of the three VR tasks were compared between groups using a two-sample t-test. For data presenting unequal variances, Welch’s version of the test was used. To determine if there was a change in VR attitude score across all participants following the VR experience, a Wilcoxon signed rank test was run (on the medians) with the alternative hypothesis that the true location is not equal to zero (median ≠ 0) which would indicate a non-zero change in attitude score. A Wilcoxon signed rank test was also conducted on VIMS medians to determine if they exceeded minimal levels after the VR experience, with the alternative hypothesis that the true median is greater than 1, which would indicate VIMS levels reported as “slightly” or above (Kim *et al*., 2018). Wilcoxon rank sum tests were run for comparisons between groups for both VR attitude change and reported VIMS. Medians (and thus the nonparametric Wilcoxon tests) were used with the non-normally distributed zero-inflated data that was expected. Analysis scripts in the R programming language along with all data are freely available at OSF: https://osf.io/45kad/?view_only=cc0f6a49e7dc49ae9af0392fb03262f0

## Results

Sixty young adults without disability participated in the simulated VR rehabilitation exercises, either with or without motor amplification. We sought to generate a young healthy reference dataset of user performance, measure attitudes towards the technology and how they change with VR exposure, gather feedback on user experience including VIMS symptoms, and identify any effect of amplification on these metrics.

Participants ranged in age from 18 to 39 years old (mean age = 19.7 years, SD = 3.22 years), with 47 participants identifying as women, 10 as men and 3 as non-binary. Three self-reported as left-handed. Anecdotally, no participants reported noticing the amplification; the use of amplification was discussed with participants at the end of participation.

During the calibration task, a Welch two-sample t-test indicated that the range of motion (ROM) for the left hand (μ = 0.41m^2^, 95% CI [0.36, 0.46], range: 0.13 – 1.17 m^2^) was not statistically different (t(117.97) = −0.266, p = 0.79) from the right-hand ROM (μ = 0.42m^2^, 95% CI [0.37, 0.47], range: 0.12 – 1.07 m^2^) (Fig. 2, Supplementary Fig. 1). No difference between hands was observed for the total distance moved during the calibration task and bubble pop game (Supplementary Fig. 2).

**Figure 2.**
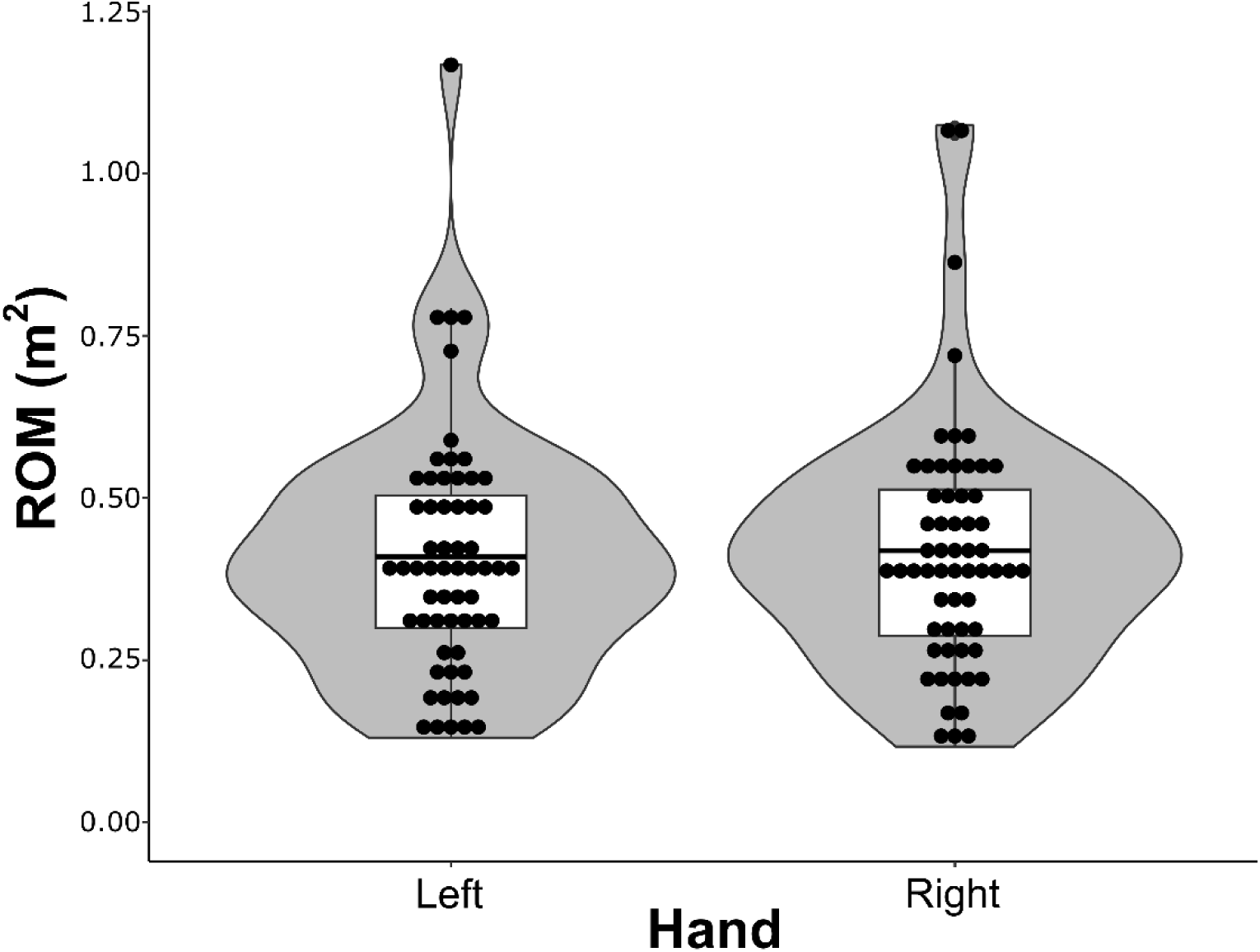
Range of motion (ROM) by hand during calibration. See Supplementary Fig. 1 for a breakdown of ROM by hand and amplification status.

Based on an individual’s difference in ROM between the hands, an amplification factor was calculated to make these ROMs equal. The amplification factor calculated for participants with smaller left-hand ROMs (μ = 1.056, 95% CI [1.034, 1.078], range: 1.003 – 1.201, n = 25) was slightly larger than those with smaller right-hand ROMs (μ = 1.035, 95% CI [1.023, 1.047], range: 1.002 – 1.164, n = 35), but this was not a statistically significant difference (t(38.19) = 1.77, p = 0.085, 95% CI [−0.0031, 0.045], Welch’s t-test) (Fig. 3). After centering the amplification factors around zero (i.e. >0 indicates a right-hand amplification), a one-sample t-test indicated the amplification factors were not statistically different from zero (μ = −0.003, t(59) = −0.376, p = 0.71), indicating no hand side bias in detected ROM in this able-bodied participant sample. Due to the nonclinical study participant population, amplification applied was likely less than would be expected in stroke survivors or other clinical populations.

**Figure 3.**
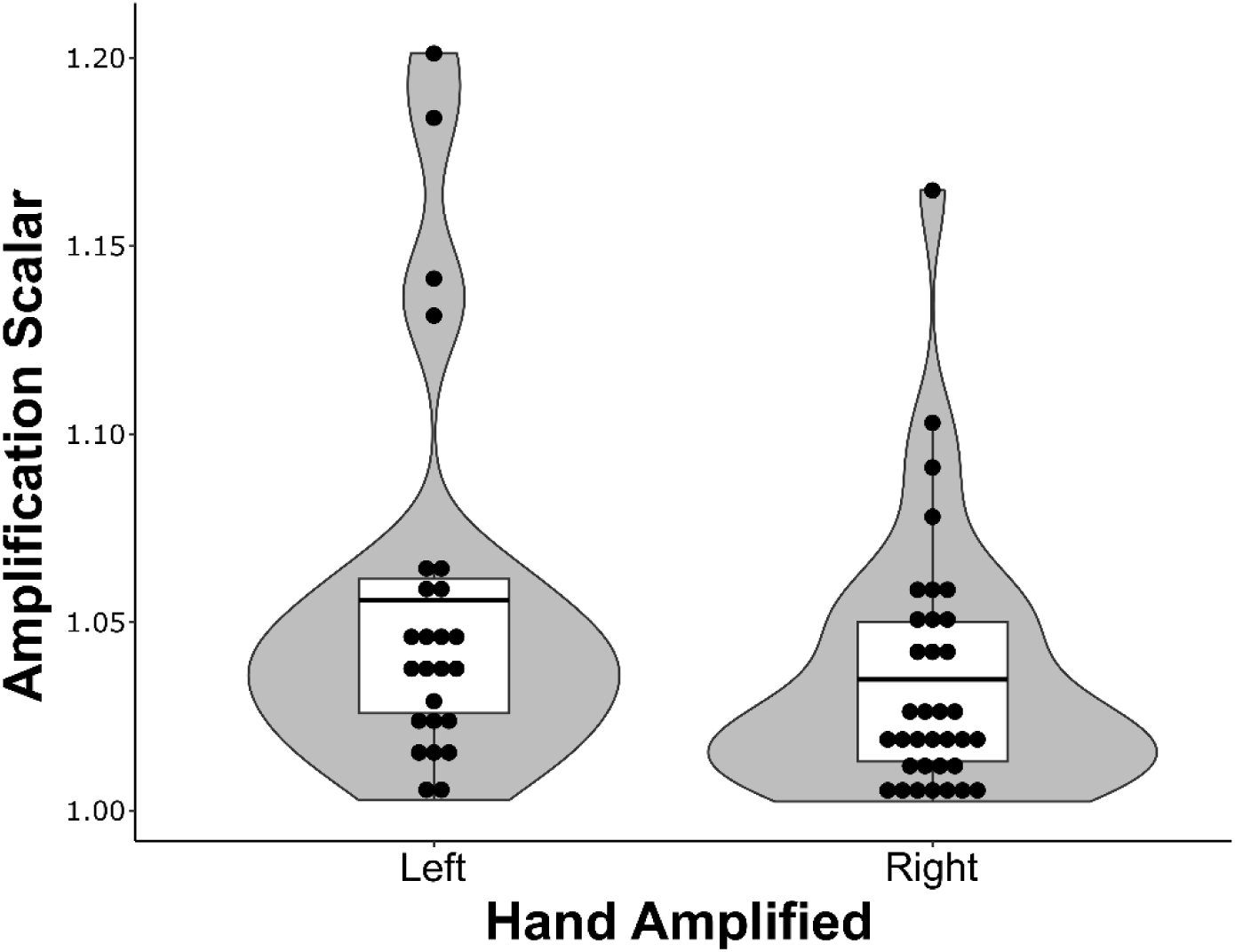
Computed amplification factor for left and right hands across all participants.

With these small amplification factors, we observed no statistically significant effect of amplification on motor performance indicators across the three games tested (Figs. 4–6). In the Bubble Pop task, the amplified participant group popped a mean of 116.4 bubbles in one minute [SD = 66.3, 95% CI [91.7, 141.2], range: 10 - 299] and the control group popped 102.5 bubbles [SD = 44.9, 95% CI [85.7, 119.3], range: 13 - 235] (Fig. 4). A Welch’s t-test indicated this was not a statistically significant difference (t(50.99) = −0.95, p = 0.34, 95% CI [−43.3, 15.4]).

**Figure 4.**
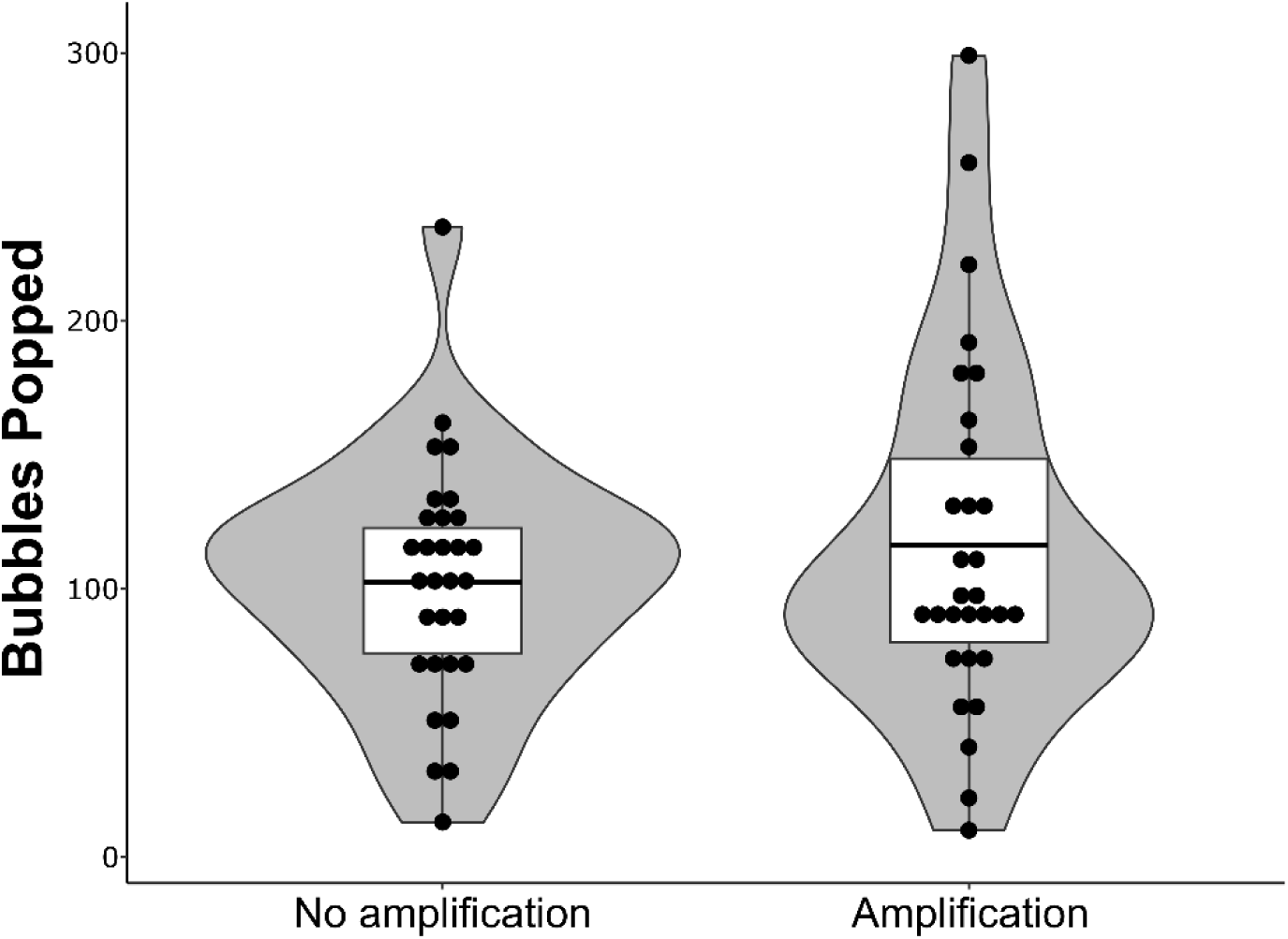
Bubble pop results for participants without and with amplification.

In the Pong Game, the amplified participant group hit the ball a mean of 26.8 times in 2 minutes [SD = 3.00, 95% CI [25.7, 28.0], range: 19 - 33] and the control group hit the ball 27.1 times [SD = 2.30, 95% CI [26.2, 28.0], range: 23 - 32] (Fig. 5). A Welch’s t-test indicated this was not a statistically significant difference (t(54.32) = 0.39, p = 0.70, 95% CI [−1.1, 1.6]).

**Figure 5.**
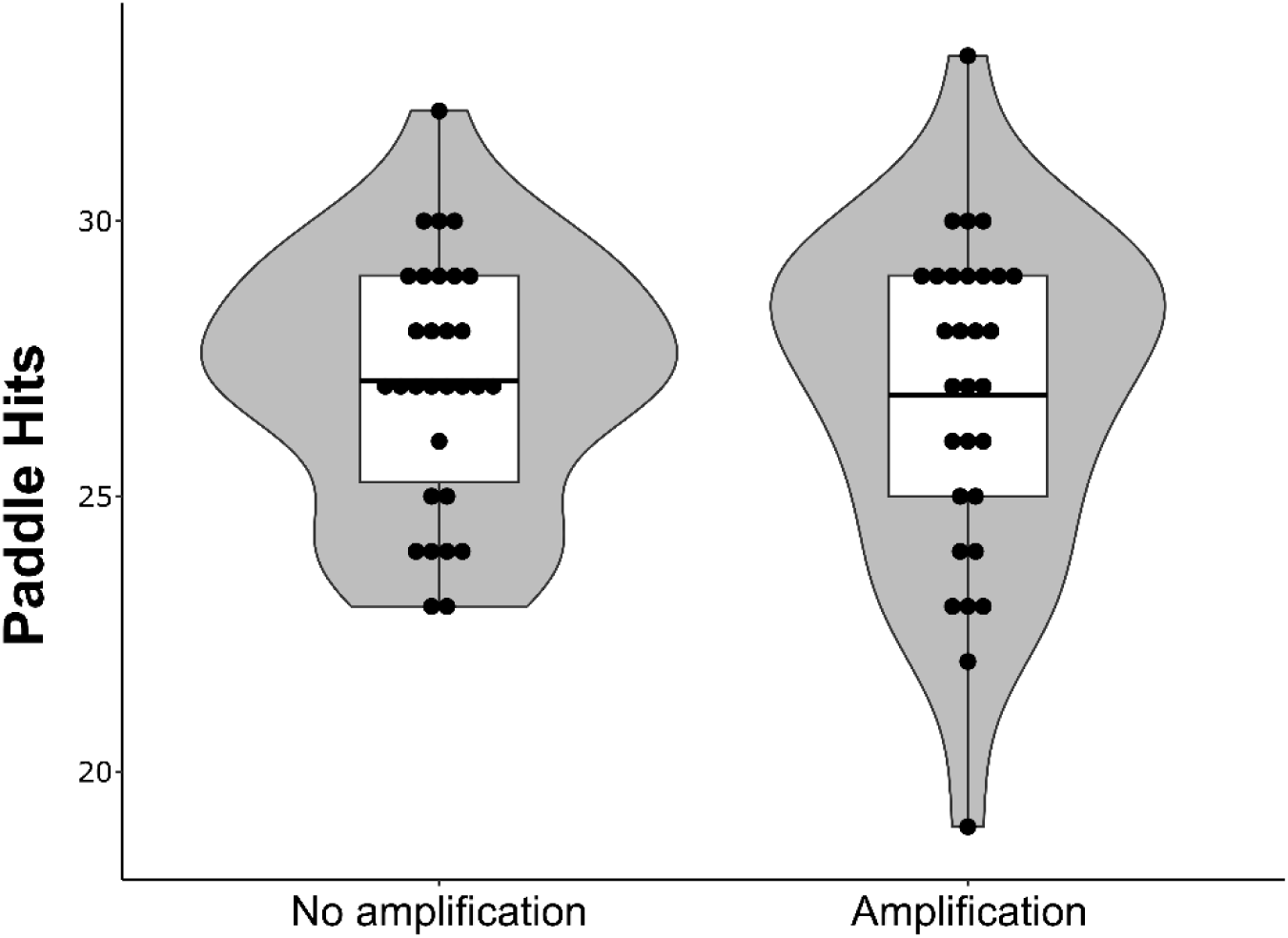
Pong Game results for participants without and with amplification.

In the Animal Feeding Task, the amplified participant group pushed a mean of 38.1 cans in two minutes [SD = 21.1, 95% CI [30.2, 45.9], range: 6 - 68] and the control group pushed 39.8 cans [SD = 18.8, 95% CI [32.8, 46.8], range: 2 - 73] (Fig. 6). A Welch’s t-test indicated this was not a statistically significant difference (t(57.24) = 0.37, p = 0.74, 95% CI [−8.6, 12.1]).

**Figure 6.**
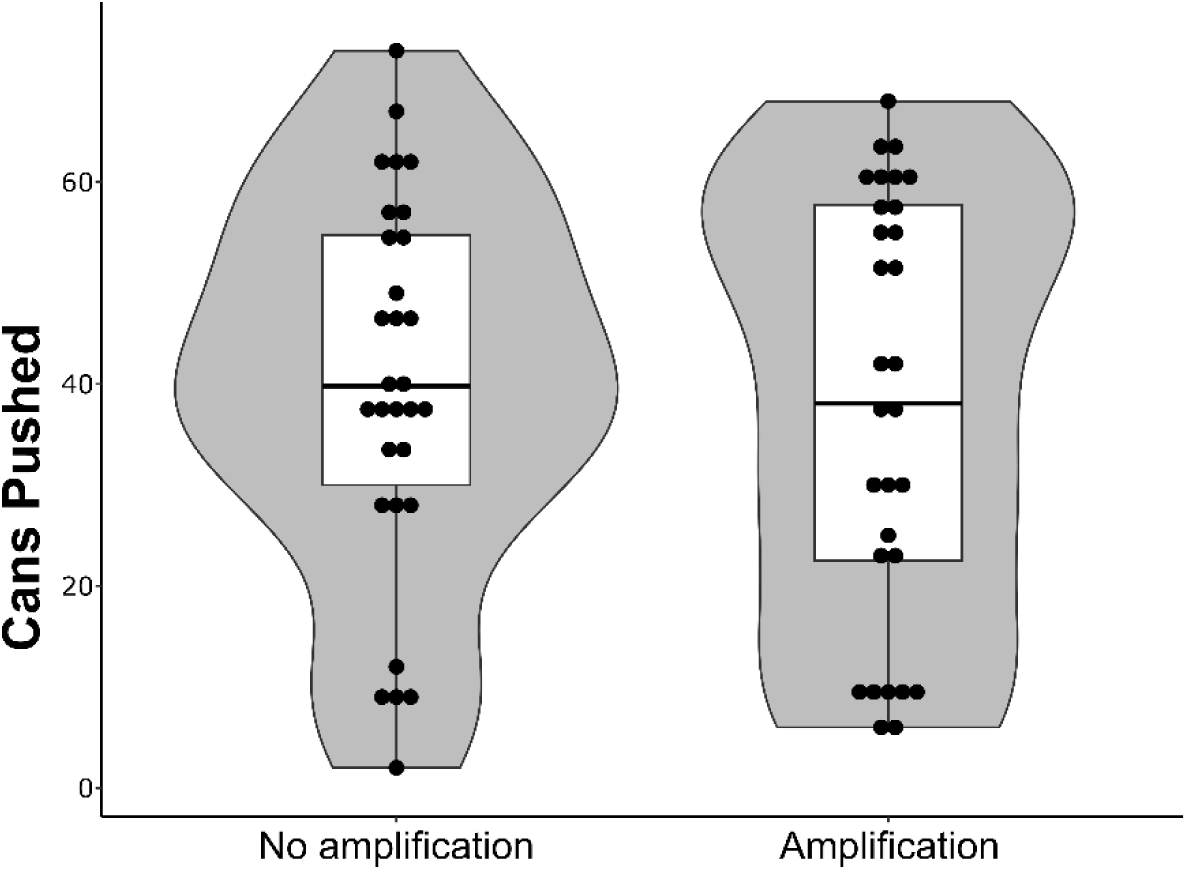
Animal Feeding results for participants without and with amplification.

Across all participants in both conditions, there was an average increase in attitude towards VR of 0.23 (SD = 0.54, 95% CI [0.094, 0.37]) (Fig. 7A), representing a small effect size (Cohen’s d = 0.44, 95% CI [0.074, 0.81]) that was statistically significant as indicated by a Wilcoxon signed rank test with continuity correction (V = 987, p < .001). The mean increases for both conditions were numerically equivalent at 0.23 per question change in VR attitude score (amplification group SD = 0.53, 95% CI [0.036, 0.43]; control group SD = 0.56, 95% CI [0.024, 0.44]) (Fig. 7B).

**Figure 7.**
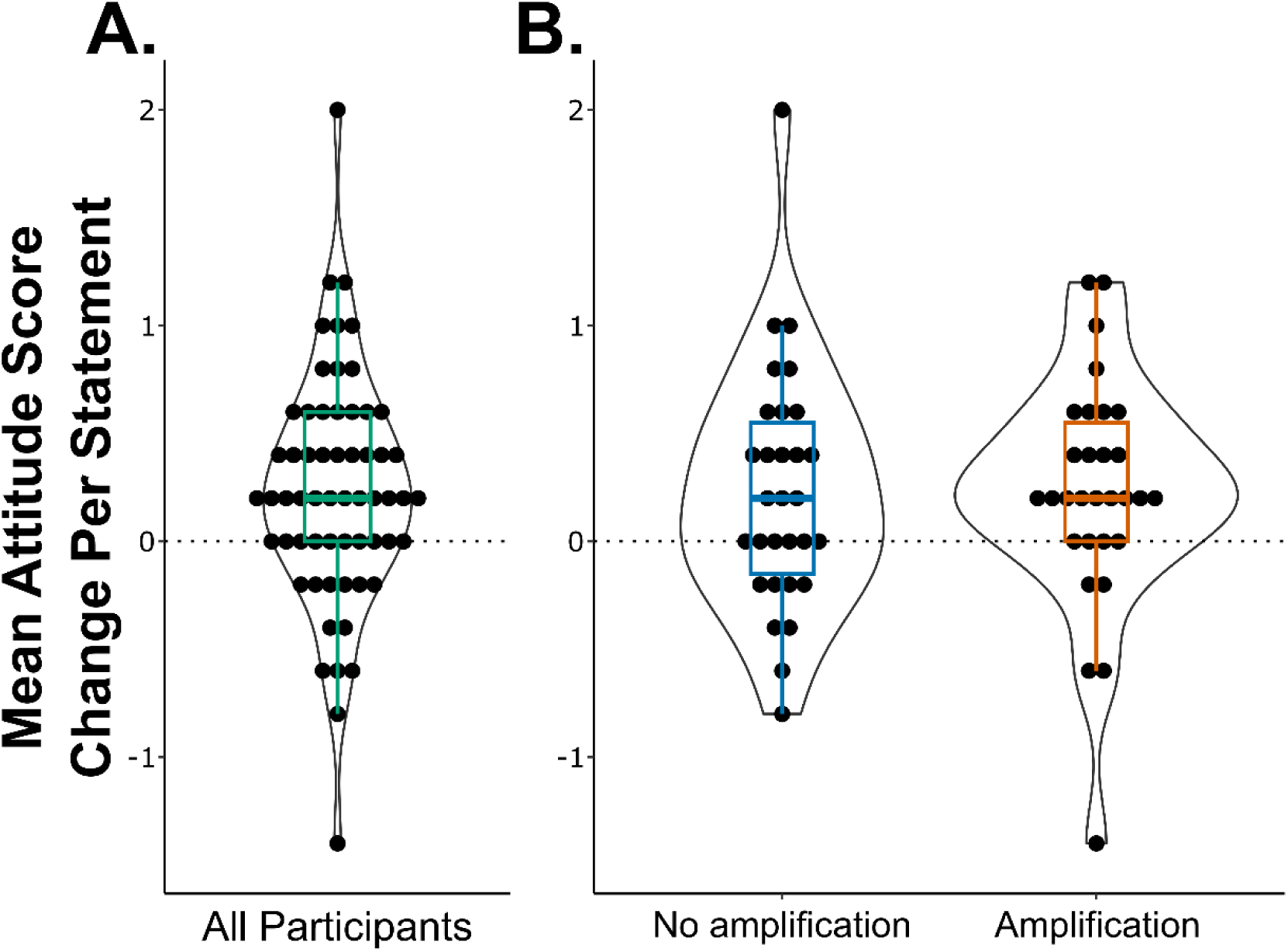
Attitude change towards VR following the VR experience. A. Mean per question score change from the pre- to post-survey for all participants. A Wilcoxon signed rank test with continuity correction showed this change to be statistically significant (V = 987, p < .001). B. Same data as in A broken down by intervention group, with and without motor amplification.

Immediately following the VR experience there was a mean VIMS score of 0.57 (SD = 0.44, 95% CI [0.45, 0.68], median = 0.4) across all participants (Fig. 8A). Amplified participants had a mean VIMS score of 0.64 (SD = 0.46, 95% CI [0.46, 0.82], median = 0.6) and non-amplified participants had a mean of 0.50 (SD = 0.41, 95% CI [0.34, 0.65], median = 0.4) participants (Fig. 8B). Eyestrain was the highest reported symptom in each group (mean for amplified = 0.93, mean for non-amplified = 0.93), followed by focus (mean for amplified = 0.83, mean for non-amplified = 0.63). On this scale, where users report the prevalence of symptoms with a zero aligning with ‘not at all’ and 4 indicating “very” as in Kim *et al*., 2018, this median score of 0.4 represents a minimal level of discomfort, well below the threshold of 1 (‘slightly’) that we tested against. Three participants did not answer all questions on the VIMS survey and their data was removed from the analysis. Of these three participants, one responded to only one question with a score of 1 (“slightly”) and the other two each skipped one question and had average scores across the answered questions of 0.125 and 1.25. A Wilcoxon signed rank test with continuity correction was run with the alternative hypothesis of u>1 given that scores above 1 would indicate more than a minor effect (“slightly” or greater VIMS symptoms). This test did not show a statistically significant effect on VIMS (V = 95.5, p = 1). A Wilcoxon rank sum test with continuity correction indicated that the difference in VIMS scores between amplified and non-amplified participants was not statistically different (W = 326, p = .20) (Fig. 8B).

**Figure 8.**
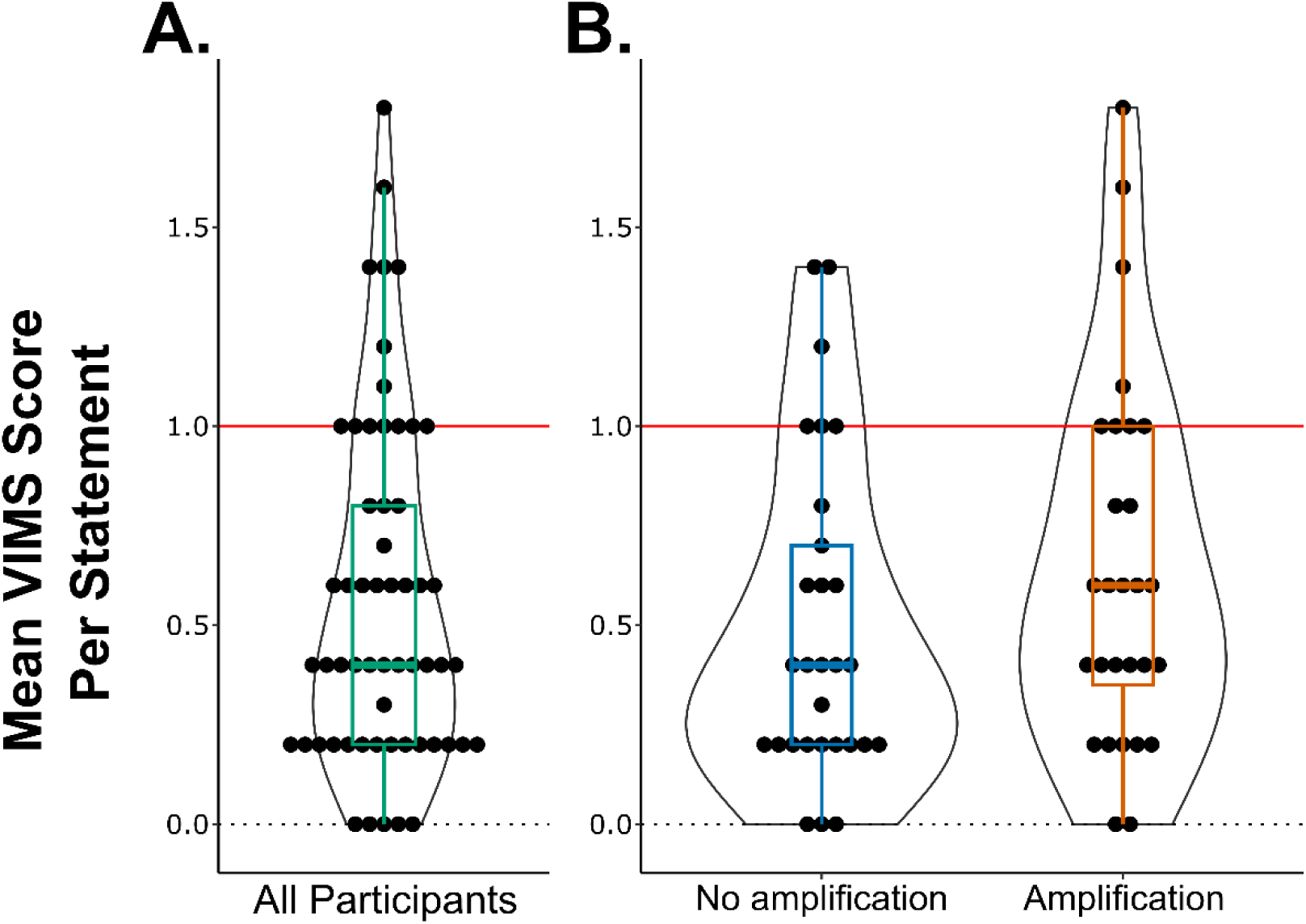
Visually induced motion sickness scores immediately following the VR experience. A. Mean score per statement for all participants. A Wilcoxon signed rank test with continuity correction did not show a statistically significant effect (u > 1; V = 95.5, p = 1). B. Same data as in A broken down by intervention group, with and without motor amplification [adapted from (Fisk, A. *et al*., 2025)].

## Discussion

We tested a virtual reality stroke rehabilitation platform in healthy young adults to assess the feasibility of its use, including its technical feasibility, initial tolerability, and acceptance, in addition to generating a young healthy reference dataset of motor performance data. Importantly, we show that motor amplification on a controller-free low-cost system has limited adverse or detrimental impacts on motor performance and VIMS. Additionally, a short exposure to the system positively influences user attitudes towards VR technology. These findings confirm the technical feasibility of applying personalized levels of motor amplification, and its initial tolerability with healthy participants which will inform future efficacy testing with stroke patients.

Importantly, we observed no statistically significant differences in motor performance between the amplification and no amplification groups. Given that the amplification introduces a visuomotor disruption, a negative impact on performance might have been expected. Our findings confirm that automated and smoothed visual amplification on a low-cost VR headset does not inadvertently degrade motor performance in healthy users. This is an important validation step to demonstrate its initial feasibility, though it is expected that future efficacy testing will employ larger amplification factors due to reduced motor function post-stroke.

Another important finding is that minimal VIMS was reported, though slightly higher VIMS levels were observed in the motor amplification group. This trend, possibly explained by sensory conflict theory, was not statistically significant but could foretell potential issues when greater amplification factors are used, such as in clinical populations. Greater amplification factors would result in a larger mismatch between visual and proprioceptive feedback, potentially exacerbating VIMS levels experienced by users. This highlights the need to monitor VIMS symptoms closely when this methodology is subsequently tested with clinical populations.

Attitudes towards technology are important predictors of active and continued engagement in rehabilitation activities (Cornet *et al*., 2019; Perski and Short, 2021) and are therefore key in early design feasibility testing. In this study, participant attitudes towards VR technology improved following the VR experience, with similar positive shifts observed in both the amplification and no amplification groups. These results mirror those of other VR studies that note positive user experiences (Mäkinen *et al*., 2022; Shang and Alena, 2025). These results are key for the continued development and testing of motor amplification within VR rehabilitation, with user acceptance playing a key role in its feasibility and future use.

As an initial feasibility assessment of this new technology, our study has several limitations that should be addressed in future work. The focus here on a healthy young adult population, while necessary, restricts the generalizability of our findings to inform the system’s future clinical deployment. A forthcoming study testing healthy older adults as well as ongoing work with stroke patients will address this limitation. Additionally, the VR exposure in this study was limited to three tasks in a single session; longer sessions or multiple sessions could induce more VIMS symptoms. However, VIMS symptoms have been shown to occur quite quickly, with one study reporting an average nausea onset time of 62 seconds (Mazloumi Gavgani *et al*., 2018). Nevertheless, ongoing comprehensive testing is needed to assess the system’s feasibility when implemented across multiple sessions and in clinical contexts. Furthermore, we did not administer a pre-session sickness questionnaire, meaning any post-session sickness scores could be confounded by pre-existing symptoms, though the low post-session scores hold value on their own. Future studies should include a pre-test to isolate the symptoms induced by the VR experience itself. Finally, small amplification factors were used among this young healthy adult sample due to a lack of motor dysfunction. Since the motor amplification is automated and based on each participant’s calibration, a healthy young adult sample would typically see minimal amplification applied to their non-dominant side. However, when testing with stroke survivors in the future, it is expected that larger motor amplification will be applied to their affected limb. This personalized level of rehabilitation support will require further testing with clinical populations to determine the impact of larger amplification on tolerability and acceptance. However, though small, the current study’s amplification demonstrates the technical feasibility of a personalized level of motor amplification.

Overall, the VR system presented here, delivering upper limb exercise with motor amplification, shows initial technical feasibility, tolerability, and acceptance among healthy young adults. Importantly, the use of a low-cost consumer headset could make this emerging treatment paradigm accessible to more patients than previous VR stroke rehabilitation systems. Before clinical deployment, next steps include a forthcoming feasibility study with healthy older adults and clinical studies with stroke survivors.

## Data Availability

All data produced are available online at Open Science Framework.

https://osf.io/45kad

---

**Supplementary Figure 1.**
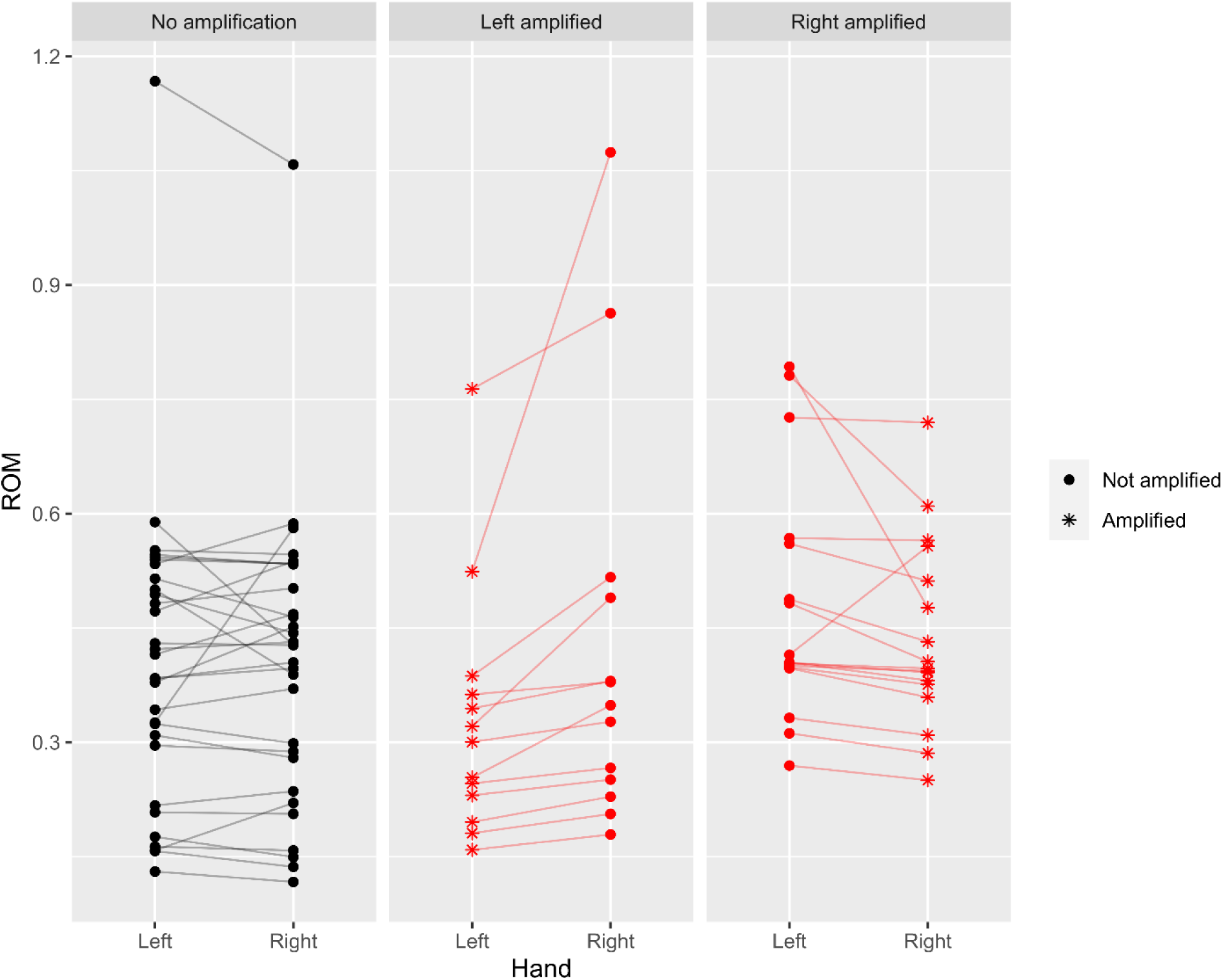
ROM calculated during calibration, broken down by hand and amplification status. Lines connect the left and right ROM for each individual participant. For amplified participants, the hand which had the lower extent in at least 2 of the 3 dimensions was selected to be amplified. For one right hand amplified participant (right panel), even with this criterion, the volume of the range of motion envelope was larger in the amplified side.

**Supplementary Figure 2.**
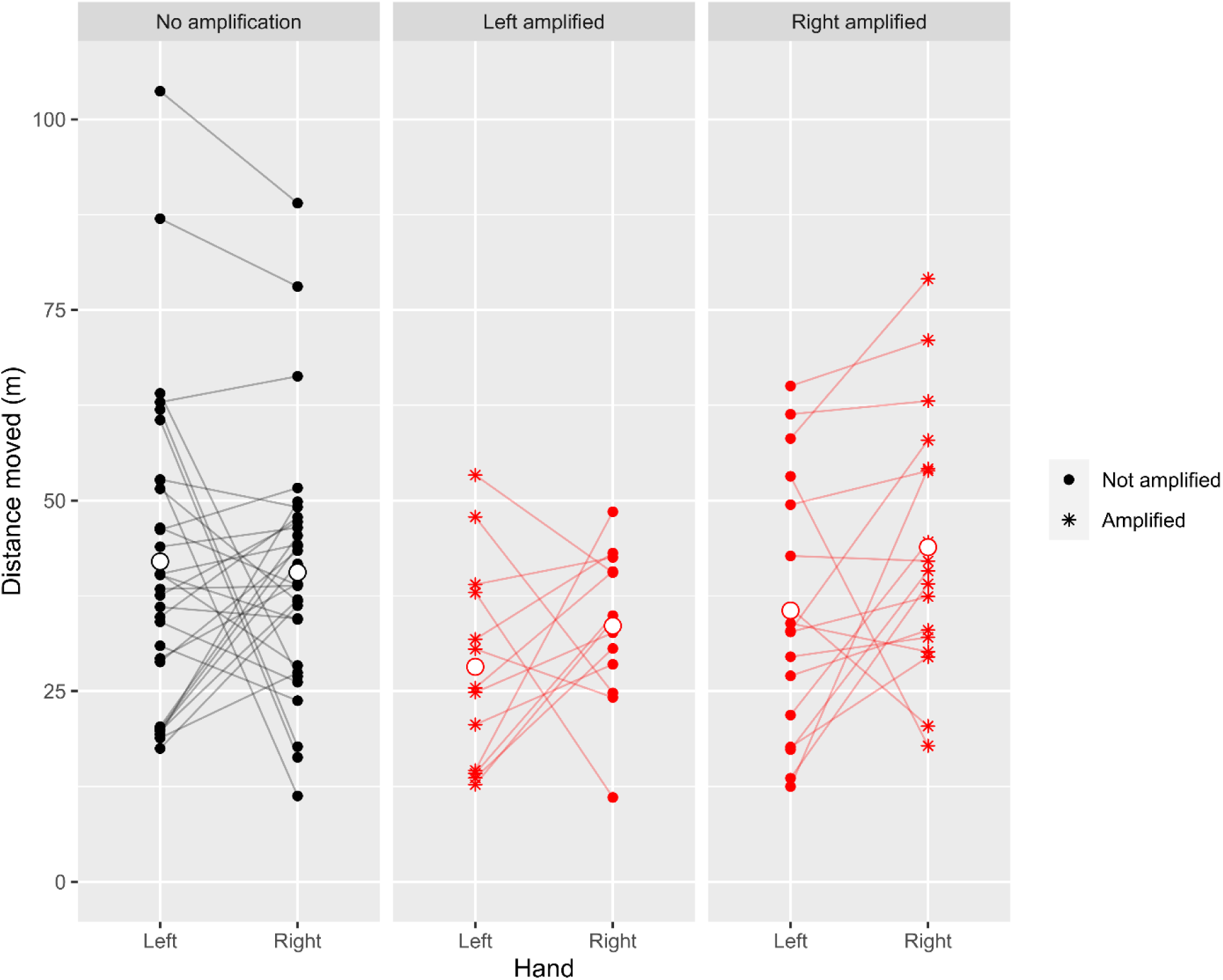
Total distance moved during the calibration phase and bubble pop task, broken down by hand and amplification status. White filled circles indicate group means.

## Notes

**Author Note** This work was supported by ResearchNS under the New Health Investigator Grant program. The authors report there are no competing interests to declare. This study was approved by the Acadia University Research Ethics Board (REB File #22-07). We thank Joy Ndirangu and Logan VanOostrum for Unity coding assistance and Colin Berkley for text editing. All data and analysis code are freely available at OSF (https://osf.io/45kad/?view_only=cc0f6a49e7dc49ae9af0392fb03262f0).

### Competing Interest Statement

The authors have declared no competing interest.

### Author Declarations

The Research Ethics Board of Acadia University (Wolfville, NS, Canada) gave ethical approval for this work (REB File # 22-07).

### Summary of Updates

Added additional descriptive statistics and emphasized the early stage feasibility focus of the work by constraining the clinical claims made.

